# The First Consecutive 5000 Patients with COVID-19 in Qatar; a Nation-wide Cohort Study

**DOI:** 10.1101/2020.07.15.20154690

**Authors:** Ali S. Omrani, Muna A. Almaslamani, Joanne Daghfal, Rand A. Alattar, Mohamed Elgara, Shahd H. Shaar, Tawheeda B. H. Ibrahim, Ahmed Zaqout, Dana Bakdach, Abdelrauof M. Akkari, Anas Baiou, Bassem Alhariri, Reem Elajez, Ahmed A. M. Husain, Mohamed N. Badawi, Fatma Ben Abid, Sulieman Abu Jarir, Shiema Abdalla, Anvar Kaleeckal, Kris Choda, Venkateswara R. Chinta, Mohamed A. Sherbash, Khalil Al-Ismail, Mohammed Abukhattab, Ali Ait Hssain, Peter V. Coyle, Roberto Bertollini, Michael P. Frenneaux, Abdullatif Al Khal, Hanan M. Al kuwari

## Abstract

**Background:** There are limited data on Coronavirus Disease 2019 (COVID-19) outcomes at a national level, and none after 60 days of follow up. The aim of this study was to describe national, 60-day all-cause mortality associated with COVID-19, and to identify risk factors associated with admission to an intensive care unit (ICU).

**Methods:** This was a retrospective cohort study including the first consecutive 5000 patients with COVID-19 in Qatar who completed 60 days of follow up by June 17, 2020. Outcomes included all-cause mortality at 60 days after COVID-19 diagnosis, and risk factors for admission to ICU.

**Results:** Included patients were diagnosed with COVID-19 between February 28 and April 17, 2020. The majority (4436, 88.7%) were males and the median age was 35 years [interquartile range (IQR) 28– 43]. By 60 days after COVID-19 diagnosis, 14 patients (0.28%) had died, 10 (0.2%) were still in hospital, and two (0.04%) were still in ICU. Fatal COVID-19 cases had a median age of 59.5 years (IQR 55.8–68), and were mostly males (13, 92.9%). All included pregnant women (26, 0.5%), children (131, 2.6%), and healthcare workers (135, 2.7%) were alive and not hospitalized at the end of follow up.

A total of 1424 patients (28.5%) required hospitalization, out of which 108 (7.6%) were admitted to ICU. Most frequent co-morbidities in hospitalized adults were diabetes (23.2%), and hypertension (20.7%). Multivariable logistic regression showed that older age [adjusted odds ratio (aOR) 1.041, 95% confidence interval (CI) 1.022–1.061 per year increase; P <0.001], male sex (aOR 4.375, 95% CI 1.964–9.744; P <0.001), diabetes (aOR 1.698, 95% CI 1.050–2.746; P 0.031), chronic kidney disease (aOR 3.590, 95% CI 1.596–8.079, P 0.002), and higher BMI (aOR 1.067, 95% CI 1.027–1.108 per unit increase; P 0.001), were all independently associated with increased risk of ICU admission.

**Conclusions:** In a relatively younger national cohort with a low co-morbidity burden, COVID-19 was associated with low all-cause mortality. Independent risk factors for ICU admission included older age, male sex, higher BMI, and co-existing diabetes or chronic kidney disease.

## 1. Introduction

Severe Acute Respiratory Syndrome Coronavirus 2 (SARS-CoV-2), the cause of Coronavirus Disease 2019 (COVID-19), emerged in China in late 2019 and has since resulted in more than 12 million confirmed infections worldwide and over 500 thousand associated deaths [1].

Based on the number of deaths as a proportion of reported COVID-19 cases, the overall estimated COVID-19-associated mortality rate is around 5.7% [2]. However, the accuracy of such a figure is uncertain given the variation in case finding policies from one healthcare setting to another [3,4]. Furthermore, reported mortality has been mostly based on in-hospital outcomes or relatively short follow up [5-9]. In their recently published recommendations for a minimal common outcome measure set for COVID-19 research, the World Health Organization (WHO) favored that mortality outcomes are assessed at 60 days [10].

Single and multi-center cohort studies suggested that risk factors for severe COVID-19 include male sex, older age, and the presence of multiple comorbidities [7,9,11]. The extent to which such risk factors are important at a population level in settings with ample healthcare resources, a COVID-19 control program based on active case finding and isolation, and a low burden of comorbidities, is unknown.

In this study, we describe 60-day outcomes of a nationwide COVID-19 cohort from Qatar, and explore patient characteristics associated with the need for admission to an intensive care unit (ICU).

## 2. Methods

Hamad Medical Corporation (HMC) encompasses multiple hospital facilities and provides all COVID-19 medical care for the 2.8 million population of Qatar. In response to the COVID-19 pandemic, existing clinical services were re-organized and two brand new hospital facilities were opened ahead of their originally planned dates. In total, non-ICU bed capacity was increased from 2,143 to 3,469 (61.9% increase), and ICU beds from 130 to 529 (306.9% increase). From a healthcare delivery perspective, HMC defines adults as those aged above 14 years.

SARS-CoV-2 infection was diagnosed by polymerase chain reaction (PCR) assays TaqPath COVID-19 Combo Kit (Thermo Fisher Scientific, Waltham, Massachusetts) or Cobas SARS-CoV-2 Test (Roche Diagnostics, Rotkreuz, Switzerland) on respiratory tract specimens. Severity of COVID-19 was classified according to the World Health Organization (WHO) guidelines [11]. Patients with asymptomatic SARS-CoV-2 infection or mild COVID-19 without significant co-morbidities were isolated in dedicated community facilities until they had two consecutive negative SARS-CoV-2 PCR results from upper airway samples taken more than 24 hours apart. COVID-19 patients with significant co-morbidities or moderate to severe disease were hospitalized for inpatient management. Standard care for hospitalized patients involved supportive care and investigational antiviral therapy. Individual regimens were selected by the treating physicians based on severity of disease, the presence of contra-indications or potential drug-drug interactions, and the patients’ preferences.

We used the HMC COVID-19 database to identify the first consecutive 5,000 patients with PCR-confirmed COVID-19 who would complete 60 days of follow up from date of diagnosis by June 17, 2020. During the period between May 24 and June 18, 2020, clinical and laboratory data were retrieved HMC’s electronic healthcare system. Final status 60 days after COVID-19 diagnosis was ascertained against the electronic healthcare system and Qatar’s national deaths records.

The primary endpoint was all-cause mortality within 60 days after RT-PCR confirmation of SARS-CoV-2 infection. For hospitalized patients, we also assessed risk factors for admission to ICU.

We summarized categorical data as numbers and percentages and compared them using Pearson’s chi-squared or Fisher’s exact test, as appropriate. Continuous data are presented as medians and interquartile ranges (IQR) and compared using Wilcoxon rank-sum test. The majority (82 patients, 75.2%) of admissions to ICU occurred within of the first 48 hours from hospitalization. We therefore used logistic regression to explore predictors of admission to ICU.

Baseline variables were included in the univariable logistic regression analysis if their between groups differences were associated with P values of <0.05. Independent variables in the multivariable regression model were chosen based on their association with P values of <0.1 in the univariable logistic regression, and on their ready availability before any COVID-19-related clinical evaluation. Due to the number of events in the study, we limited the number of independent variables in the multivariable regression analysis to eight to avoid overfitting the model. The final multivariable logistic regression model included age, male sex, body mass index (BMI), defined as body weight in kilograms divided by squared height in meters, and co-existing diabetes mellitus, systemic hypertension, coronary artery disease, chronic liver disease, and chronic kidney disease. Multiple imputations approach was applied for variables with >5% missingness.

All P values were two-sided with a threshold of <0.05 for statistical significance. Statistical analyses were performed using Stata Statistical Software Release 15.1 (StataCorp LLC, College Station, Texas). The report was prepared according to STROBE recommendations [12]. The study was approved by HMC’s Institutional Review Board (MRC0120191), with a waiver of informed consent.

## 3. Results

Individuals included in this study were diagnosed with SARS-CoV-2 infection between February 28 and April 17, 2020. Initial SARS-CoV-2 cases were diagnosed in travelers returning from Iran and Europe. Sustained local transmission became established thereafter (Figure 1). Of the 5,000 PCR-confirmed COVID-19 cases included in this report, 4,436 (88.7%) were in males and the majority belonged to age groups 25–34 years (1811, 36.2%) and 35–44 years (1445, 28.9%) (Figure 2). The cohort included 131 (2.6%) individuals aged 14 years or less, 26 (0.5%) pregnant women and 135 (2.7%) healthcare workers (Table S1 in the Supplementary material).

**Figure 1.**
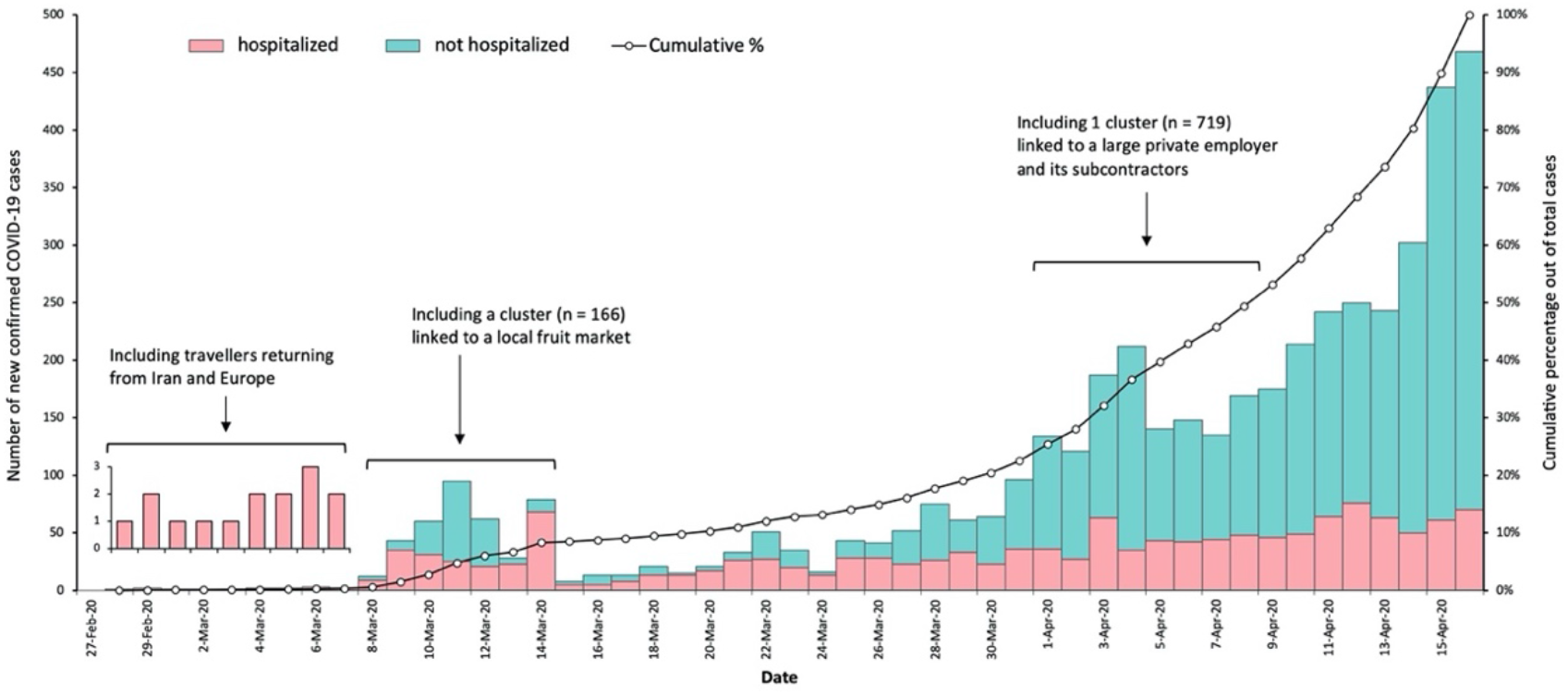
Epidemic Curve of the Cases of Coronavirus Disease 2019 (COVID-19) in Qatar. Daily numbers (Y axis) and cumulative count (Z axis) of confirmed cases are plotted by date (X axis) samples were taken. Inserts describe significant events with their corresponding dates. A total of 474 cases were diagnosed on April 17, 2020, of which 251 are included in this report.

**Figure 2.**
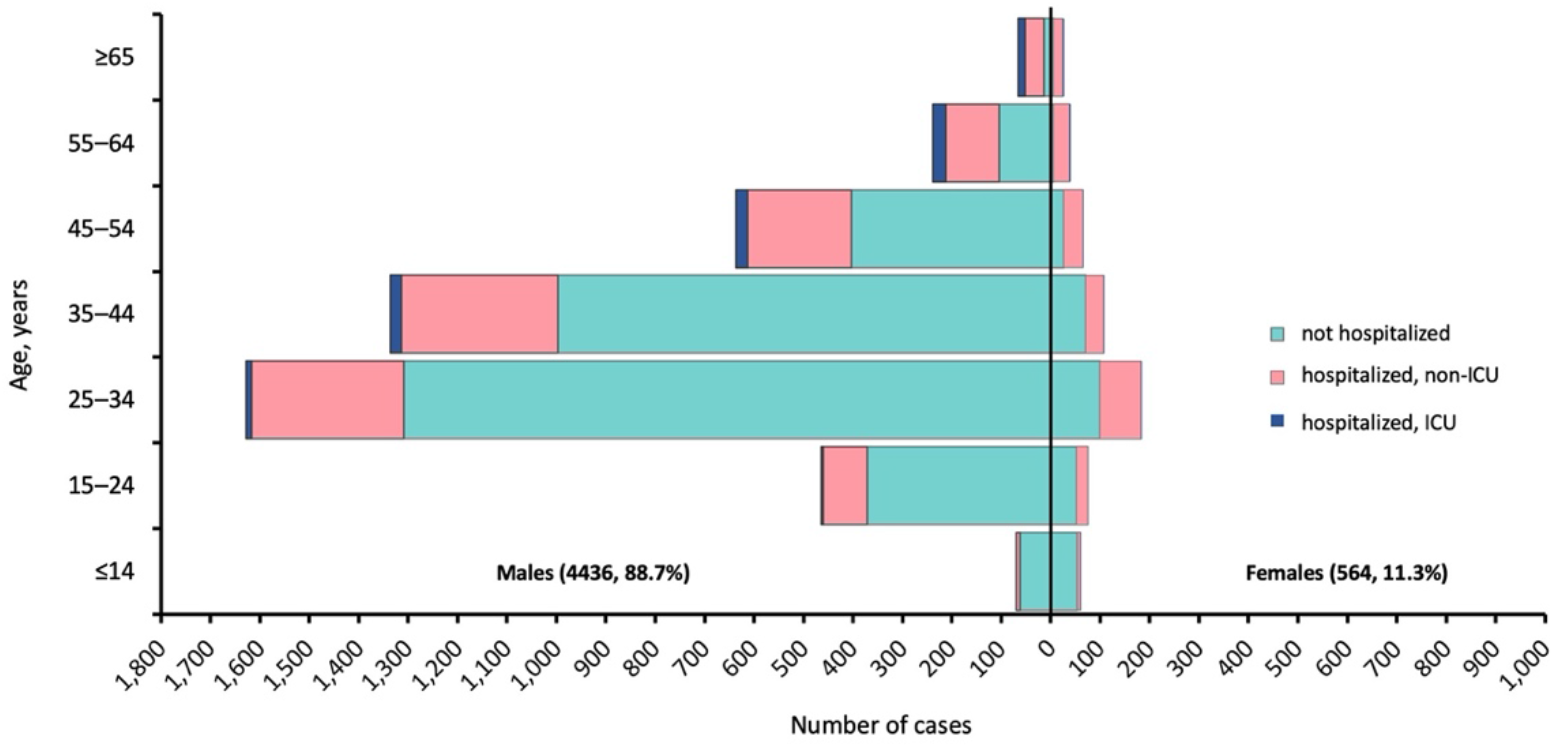
Patients with Coronavirus Disease 2019 (COVID-19) Stratified by Age Group and Sex X axis, numbers by sex; Y axis age group (years)

The majority of individuals in this study did not require hospitalization (3,576, 71.5%). Of 1,424 patients who required hospitalization, 108 (7.6%) were admitted to ICU. Overall, 60 days after COVID-19 diagnosis, 14 patients (0.28%) had died, 10 patients (0.2%) were still in hospital and two (0.04%) were still in ICU (Figure 3).

**Figure 3.**
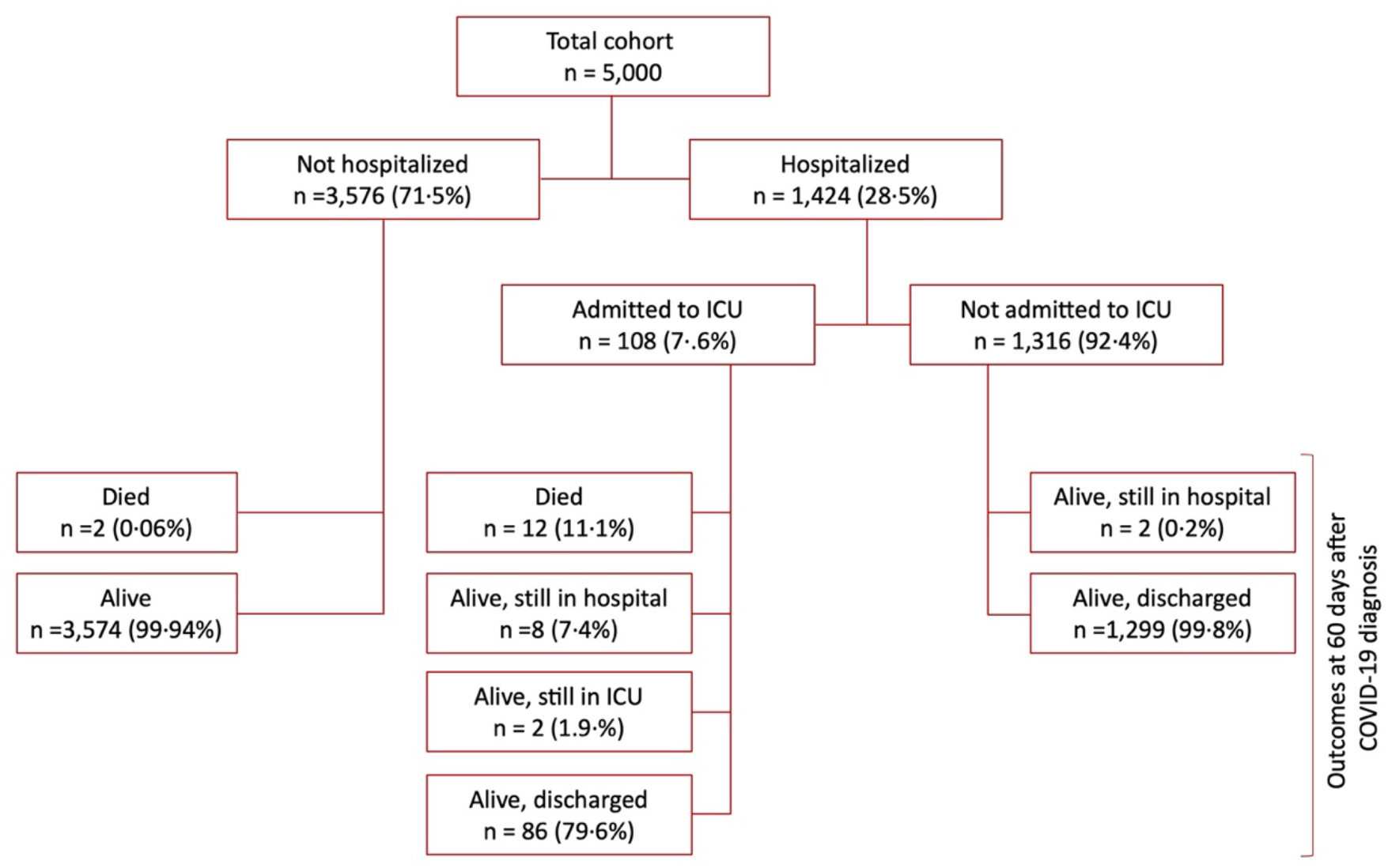
60 Day Outcomes of 5000 Patients with Coronavirus Disease 2019 (COVID-19)

### 3.1. Hospitalized Adults

Out of 4,869 individuals aged >14 included in this report, 1409 (28.9%) were hospitalized. The majority (1167, 82.8%) were males and the median age was 39 years (IQR 30–50). Nationalities from WHO’s South-East Asia Region (791, 56.1%) and Eastern Mediterranean Region (485, 34.4%) were most frequent. Diabetes (327, 23.2%) and hypertension (292, 20.7%) were the most common co-existing medical conditions. Fever (58.3%) and cough (59.1%) were the most common presenting symptoms. Median BMI was 26.8 kg/m^2^ (IQR 23.9–29.8). Most patients (1233, 87.5%) did not require oxygen therapy within the first 24 hours of hospitalization. Hydroxychloroquine (1044, 74.1%), azithromycin (898, 63..7%), and lopinavir-ritonavir (522, 37%) were the most commonly used investigational antiviral agents.

Compared with those who did not require ICU admission, ICU patients were significantly more likely to be males (P 0.005), have higher median age (P <0.001) and to have multiple co-morbidities (P <0.001) (Table 1). They also had significantly higher median BMI (28.2 versus 26.6, P <0.001) and were more likely to present with fever, cough and dyspnea (P <0.001 for each). Within the first 24 hours of hospitalization, ICU patients had significantly higher median heart rate (95 versus 86 per minute, P <0.001), and respiratory rate (26.5 versus 19 per minute, P <0.001), significantly lower oxygen saturation (94% versus 98%, P <0.001) (Table 1). In addition, baseline blood investigations from ICU patient were significantly more likely to show lower median peripheral lymphocyte count (1.0 versus 1.7 ×10^9^/L, P <0.001), and higher median serum creatinine (90 versus 80 µmol/L, P <0.001), and CRP (107.7 versus 7, P <0.001). Complications such as acute kidney injury (43.5% versus 1.5%), and myocardial injury (14.8% versus 0.3%, P <0.001) were more common in ICU compared with non-ICU patients. Other baseline characteristics, management, and complications variables in hospitalized COVID-19 adults included in this study are shown in Table 1.

**Table 1.** Baseline characteristics, management and complications in individuals aged >14 years hospitalized with Coronavirus Disease 2019.

| Variable | All patients<br>(n = 1409) | Non-ICU<br>(n = 1301) | ICU<br>(n = 108) | P value |
| --- | --- | --- | --- | --- |
| **Baseline characteristics** |  |  |  |  |
| Male sex | 1167 (82.8%) | 1067 (82%) | 100 (92.6%) | 0.005 |
| Age (years) | 39 (30–50) | 38 (30–49) | 49.5 (39.5–60) | <0.001 |
| Age group (years) |  |  |  | <0.001 |
| 15–24 | 116 (8.2%) | 113 (8.7%) | 3 (2.8%) |  |
| 25–34 | 403 (28.6%) | 391 (30.1%) | 12 (11.1%) |  |
| 35–44 | 377 (26.7%) | 353 (27.1%) | 24 (22.2%) |  |
| 45–54 | 275 (19.5%) | 250 (19.2%) | 25 (23.1%) |  |
| 55–64 | 166 (11.8%) | 138 (10.6%) | 28 (25.9%) |  |
| ≥65 | 72 (5.1%) | 56 (4.3%) | 16 (14.8%) |  |
| Healthcare workers | 63 (4.5%) | 60 (4.6%) | 3 (2.8%) | 0.48 |
| **Nationality according to WHO region** |  |  |  | 0.005 |
| African Region | 29 (2.1%) | 27 (2.1%) | 2 (1.9%) |  |
| Eastern Mediterranean Region | 485 (34.4%) | 449 (34.5%) | 36 (33.3%) |  |
| European Region | 24 (1.7%) | 22 (1.7%) | 2 (1.9%) |  |
| Region of the Americas | 13 (0.9%) | 11 (0.8%) | 2 (1.9%) |  |
| South-East Asia Region | 791 (56.1%) | 739 (56.8%) | 52 (48.1%) |  |
| Western Pacific Region | 67 (4.8%) | 53 (4.1%) | 14 (13%) |  |
| **Co-morbidities** |  |  |  |  |
| Pregnant | 19 (1.3%) | 18 (1.4%) | 1 (0.9%) | 1.0 |
| Diabetes mellitus | 327 (23.2%) | 275 (21.1%) | 52 (48.1%) | <0.001 |
| Hypertension | 292 (20.7%) | 248 (19.1%) | 52 (48.1%) | <0.001 |
| Coronary artery disease | 41 (2.9%) | 31 (2.4%) | 10 (9.3%) | <0.001 |
| Chronic lung disease | 83 (5.9%) | 73 (5.6%) | 10 (9.3%) | 0.12 |
| Chronic liver disease | 18 (1.3%) | 14 (1.1%) | 4 (3.7%) | 0.043 |
| Chronic kidney disease | 34 (2.4%) | 21 (1.6%) | 13 (12%) | <0.001 |
| Current malignant diagnosis | 20 (1.4%) | 18 (1.4%) | 2 (1.9%) | 0.66 |
| Number of comorbidities |  |  |  | <0.001 |
| None | 894 (63.4%) | 859 (66.0%) | 35 (32.4%) |  |
| One comorbidity | 301 (21.4%) | 268 (20.6%) | 33 (30.6%) |  |
| Two comorbidities | 144 (10.2%) | 124 (9.5%) | 20 (18.5%) |  |
| More than two comorbidities | 70 (5.0%) | 50 (3.8%) | 20 (18.5%) |  |
| Current or past smoker | 130/858 (15.2%) | 121/806 (15%) | 9/52 (17.3%) | 0.71 |
| **Mode of presentation** |  |  |  | <0.001 |
| Screening or contact tracing | 390/1401 (27.8%) | 390/1293 (30.2%) | 0 |  |
| Symptomatic | 1,011/1401 (72.2%) | 903/1293 (69.8%) | 108 (100%) |  |
| **Symptoms** |  |  |  |  |
| Fever | 815/1397 (58.3%) | 710/1289 (55.1%) | 105 (97.2%) | <0.001 |
| Cough | 825/1397 (59.1%) | 730/1289 (56.6%) | 97 (89.8%) | <0.001 |
| Sore throat | 366/1397 (26.2%) | 345/1289 (26.8%) | 21 (19.4%) | 0.11 |
| Rhinorrhea | 131/1397(9.4%) | 130/1289 (10.1%) | 1 ( 0.9%) | <0.001 |
| Dyspnea | 211/1397 (15.1%) | 156/1289 (12.1%) | 56 (51.9%) | <0.001 |
| Fatigue | 136/1397 (9.7%) | 121/1289 ( 9.4%) | 15 (13.9%) | 0.13 |
| Generalized pain | 286/1397(20.%) | 265/1289 (19.9%) | 21 (19.4%) | 0.9 |
| Diarrhea | 55/1397 (3.9%) | 53/1289 (4.1%) | 2 (1.9%) | 0.43 |
| Nausea and/or vomiting | 62/1397 (4.2%) | 52/1289 (4.0%) | 10 (9.3%) | 0.024 |
| **Measurements, vital signs, and laboratory results within the first 24 hours of hospitalization** |  |  |  |  |
| Body mass index<sup>a</sup> (kg/m<sup>2</sup>) | 26.8 (23.9–29.8) | 26.6 (23.8–29.7) | 28.2 (25.8–31.6) | <0.001 |
| Systolic blood pressure (mmHg) | 117 (108–127) | 117.0 (108–127) | 116 (103.5–126) | 0.074 |
| Temperature<sup>b</sup> (° C) | 37.1 (36.8–38.0) | 37.0 (36.8–37.9) | 37.8 (37.1–38.7) | <0.001 |
| Hear rate<sup>c</sup> (beats per minute) | 87.0 (78.0–98.0) | 86 (78–97) | 95 (86–108) | <0.001 |
| Respiratory rate<sup>d</sup> (breaths per minute) | 19 (18–20) | 19 (18–20) | 26.5 (20–32.5) | <0.001 |
| Oxygen saturation<sup>e</sup> (%) | 97 (95–99) | 98 (96–99) | 94 (91–96) | <0.001 |
| White blood cell count<sup>f</sup> (x10<sup>9</sup> cells per L) | 6.4 (5–8) | 6.4 (5.0–7.9) | 6.6 (5.2–8.6) | 0.058 |
| Lymphocyte counts<sup>g</sup> (x10<sup>9</sup> cells per L) | 1.6 (1.1–2.2) | 1.7 (1.2–2.2) | 1.0 (0.7–1.3) | <0.001 |
| Platelet counts<sup>h</sup> (x10<sup>9</sup> cells per L) | 232 (188–280) | 235 (189–283) | 202.5 (171.5–242.5) | <0.001 |
| Serum sodium<sup>h</sup> (mmol/L) | 138 (135–140) | 138 (136–140) | 135 (133–137) | <0.001 |
| Serum creatinine<sup>i</sup> (μmol/L) | 80 (68–90) | 80 (68–90) | 90 (76.5–108.5) | <0.001 |
| CRP<sup>i</sup> (mg/L) | 8.9 (5–46) | 7.0 (5–33.8) | 107.7 (55.3–169.5) | <0.001 |
| ALT<sup>k</sup> (U/L) | 27 (19–41) | 27 (19–40) | 32 (21.5–50.5) | <0.001 |
| Chest radiology showing pulmonary infiltrates | 559/1383 (40.4%) | 457/1275 (35.8%) | 102 (94.4%) | <0.001 |
| **Management** |  |  |  |  |
| **Highest respiratory support in the first 24 hours of hospitalization** |  |  |  | <0.001 |
| Ambient air | 1,233 (87.5%) | 1,226 (94.2%) | 7 (6.5%) |  |
| Oxygen via face mask or nasal canulae | 122 (8.5%) | 75 (5.8%) | 45 (41.7%) |  |
| Non-invasive mechanical ventilation | 14 (1.0%) | 0 | 14 (13%) |  |
| Invasive mechanical ventilation | 42 (3.0%) | 0 | 42 (38.9%) |  |
| Invasive mechanical ventilation anytime during hospitalization | 91 (6.5%) | 0 | 91 (84.3%) | <0.001 |
| Vasopressor support | 68 (4.8%) | 2 ( 0.2%) | 66 (61.1%) | <0.001 |
| Renal replacement therapy | 18 (1.3%) | 4 ( 0.3%) | 16 (14.8%) | <0.001 |
| **Antiviral and Anti-inflammatory Therapy** |  |  |  |  |
| Hydroxychloroquine | 1044 (74.1%) | 936 (71.9%) | 108 (100.0%) | <0.001 |
| Azithromycin | 898 (63.7%) | 791 (60.8%) | 107 (99.1%) | <0.001 |
| Lopinavir–ritonavir | 522 (37%) | 435 (33.4%) | 87 (80.6%) | <0.001 |
| Ribavirin | 107 (7.6%) | 33 (2.5%) | 74 (68.5%) | <0.001 |
| Interferon | 27 (1.9%) | 1 ( 0.1%) | 26 (24.1%) | <0.001 |
| Tocilizumab | 111 ( 7.9%) | 12 ( 0.9%) | 99 (91.7%) | <0.001 |
| Systemic corticosteroids | 78 ( 5.5%) | 7 ( 0.5%) | 71 (65.7%) | <0.001 |
| **Complications** |  |  |  |  |
| Acute respiratory distress syndrome | 94 (6.7%) | 0 | 94 (87%) | <0.001 |
| Acute kidney injury | 67 (4.8%) | 20 (1.5%) | 47 (43.5%) | <0.001 |
| Myocardial injury | 20 (1.4%) | 4 (0.3%) | 16 (14.8%) | <0.001 |
| Thromboembolism | 3 (0.2%) | 0 | 3 (2.8%) | <0.001 |
| Arrhythmia | 4 (0.3%) | 0 | 4 (3.7%) | <0.001 |
| **Length of stay (days)** |  |  |  |  |
| Hospital length of stay | 7 (4–14) | 7 (3–12) | 24.5 (19–37.5) | <0.001 |
| ICU length of stay | 12 (8–21) | NA | 12 (8–21) | NA |

Data are median (IQR), n (%), or n/N (%), where N is the total number of patients with available data. P values comparing ICU and non-ICU are from Pearson's chi-squared test, Fisher's exact test, or Wilcoxon rank-sum test. <sup>a</sup>Data missing for 246 (17.5%). <sup>b</sup>Data missing for 16, (1.1%). <sup>c</sup>Data missing for 179 (12.7%). <sup>d</sup>Data missing for 33 (2.3%). <sup>e</sup>Data missing for 20 (1.4%). <sup>f</sup>Data missing for 37 (2.6%). <sup>g</sup>Data missing for 42 (3%). <sup>h</sup>Data missing for 39 (2.8%). <sup>i</sup>Data missing for 43 (3.1%). <sup>j</sup>Data missing for 148 (10.5%). <sup>k</sup>Data missing for 83 (5.9%). ALT, alanine transaminase; CRP, C-reactive protein; ICU, intensive care unit; NA, not applicable; WHO, World Health Organization.

### 3.2. Risk factors for ICU admission

In the univariable logistic regression analysis, the odds of admission to ICU were significantly higher in older patients, males compared with females, and in those with diabetes, hypertension, coronary artery disease, or chronic lung, liver, or kidney disease, and in those with higher BMI (Table 2). The presence of cough, dyspnea, or fever, elevated baseline heart rate or respiratory rate, decreased oxygen saturation, lower lymphocyte count, and increased serum creatinine, CRP, and ALT were also associated with admission to ICU (Table 2).

**Table 2.** Risk factors associated with admission of patients with COVID-19 to ICU

| Variable | Univariable OR<br>(95% CI, P value) | Multivariable OR<br>(95% CI, P value) |
| --- | --- | --- |
| **Demographics** |  |  |
| Age (per year increase) | 1.056 (1.041–1.071, P <0.001) | 1.041 (1.022–1.061, P <0.001) |
| Male sex (versus female) | 2.741 (1.316–5.711, P 0.007) | 4.375 (1.964–9.744, P <0.001) |
| **Co-morbidity present** |  |  |
| Diabetes mellitus | 3.461 (2.319–5.164, P <0.001) | 1.698 (1.050–2.746, P 0.031) |
| Hypertension | 2.919 (1.941–4.340, P <0.001) | 0.980 (0.589–1.630, P 0.937) |
| Coronary artery disease | 4.180 (1.991–8.778, P <0.001) | 1.090 (0.449–2.643, P 0.849) |
| Chronic lung disease | 1.717 (0.859–3.430, P 0.126) | not applicable |
| Chronic liver disease | 3.536 (1.143–10.934, P 0.028) | 2.463 (0.716–8.465, P 0.153) |
| Chronic kidney disease | 8.341 (4.050–17.177, P <0.001) | 3.590 (1.596–8.079, P 0.002) |
| BMI (per one kg/m<sup>2</sup> increase) | 1.067 (1.033–1.102, P <0.001) | 1.067 (1.027–1.108, P 0.001) |
| **Presenting symptoms** |  |  |
| Fever | 28.542 (9.011–90.401, P <0.001) | not applicable |
| Cough | 6.145 (3.339–11.312, P <0.001) | not applicable |
| Dyspnea | 7.879 (5.123–11.909, P <0.001) | not applicable |
| Sore throat | 0.660 (0.403–1.079, P <0.001) | not applicable |
| Rhinorrhea | 0.833 (0.0115–0.602, P 0.014) | not applicable |
| **Baseline vital signa (per unit increase)** |  |  |
| Systolic blood pressure (mmHg) | 0.987 (0.973–1.001, P 0.062) | not applicable |
| Heart rate (beats per minute) | 1.037 (1.024–1.049, P <0.001) | not applicable |
| Respiratory rate (breaths per minute) | 1.264 (1.217–1.313, P <0.001) | not applicable |
| Oxygen saturation (%) | 0.651 (0.605–0.701, P <0.001) | not applicable |
| **Baseline laboratory results (per unit increase)** |  |  |
| Lymphocytes count (x 10<sup>9</sup> per L) | 0.155 (0.100–0.240, P <0.001) | not applicable |
| Platelet count (x 10<sup>9</sup> per L) | 0.993 (0.990–0.996, P <0.001) | not applicable |
| Serum sodium (mmol/L) | 0.801 (0.757–0.848, P <0.001) | not applicable |
| Serum creatinine (μmol/L) | 1.003 (1.001–1.005, P <0.005) | not applicable |
| CRP (mg/L) | 1.016 (1.013–1.019, P <0.001) | not applicable |
| ALT (U/L) | 1.001 (0.999–1.002, P 0.2) | not applicable |

ALT, alanine transaminase; BMI, body mass index (calculated as weight in kilograms divided by height in meters squared); CI, confidence interval; COVID-19, coronavirus disease 2019; CRP, C-reactive protein; ICU, intensive care unit; IQR, interquartile range.; OR, odds ratio

In the multivariable model, we found that older age, male sex, co-existing diabetes or chronic kidney disease, and higher BMI were all independently associated with increased risk of need for ICU admission (Table 2).

### 3.3. Fatal COVID-19

A total of 14 patients (0.28%) died within 60 days of follow up. The median age of fatal COVID-19 cases was 59.5 years (IQR 55.8–68). The majority of deceased patients were males (13, 92.9%) and most (8, 57.1%) had two or more co-morbidities (Table S2 in the Supplementary Material).

Two patients died without hospitalization. The first was a 59 year-old man with a history of hypertension and heavy smoking. He had asymptomatic SARS-CoV-2 infection and was isolated in a community facility pending viral clearance. He developed severe chest pain and cardiopulmonary arrest 17 days after COVID-19 diagnosis. The second patient was a 74 year old man with end-stage kidney disease, hypertension, diabetes and coronary artery disease. He developed cardiopulmonary arrest shortly after presenting to the emergency department in severe respiratory distress. A post-mortem examination to confirm the cause of death was not performed in either case.

The remaining 12 deaths all occurred in patients who were in ICU with severe ARDS requiring prolonged invasive mechanical ventilation. Deaths occurred after a median of 24 days (IQR 14–49) from COVID-19 diagnosis. Ten (86.5%) deaths occurred in patients aged 55 or older. The remaining two were in patients aged 54 years and 24 years. The former had diabetes, hypertension, and obesity (BMI 38.7). The latter patient presented with fulminant hepatitis and his hepatitis B serology was positive for surface IgM antibodies. He died within 11 days with encephalopathy and multi-organ failure.

### 3.4. Pregnant women with SARS-CoV-2 infection

The study included 26 pregnant women with SARS-CoV-2 infection with median age of 29 years (IQR 25.5–33). Nineteen (73.1%) were hospitalized, including one (3.8%) in ICU, and all were discharged within the follow up period. Ten (38.5%) pregnant women with COVID-19 gave birth during the follow up period; all resulting in healthy babies with negative SARS-CoV-2 tests (Table S3 in the Supplementary Material).

### 3.5. Healthcare workers with SARS-CoV-2 infection

A total of 135 patients in this cohort were healthcare workers. Their median age was 35 years (28–43) and the majority were males (101, 74.8%). The most frequent professional background of affected healthcare workers was nursing (49, 36.3%), and allied healthcare (27, 20%) (Table S4 in The Supplementary Material). Out of 63 (46.6%) who required hospitalization, three (2.2%) required admission to ICU. All healthcare workers in this study were alive and out of hospital at end of follow up (Table S4 in the Supplementary Material).

### 3.6. Children with SARS-CoV-2 infection

There were 131 individuals aged 14 years or less in the study, of which 69 (52.7%) were males. Median age was 7 years (IQR 4–10). Children were mostly diagnosed in the context of contact screening (75/123, 61%), and were not hospitalized (116, 88.5%). The majority (120, 91.6%) of children, including all seven infants, had family members with confirmed COVID-19 (Table S5 in the Supplementary Material).

## 4. Discussion

In this national COVID-19 cohort, only 14 (0.28%) out of 5,000 patients died within 60 days of diagnosis, and 12 (0.24%) required ongoing hospitalization at the end of the 60-day follow up period.

Our mortality rates are considerably lower than previously reported form large COVID-19 cohorts from China, Europe and United States [5-9]. There are several possible explanations for our findings. Firstly, as of July 11, 2020, the total number of SARS-CoV-2 RT-PCR tests performed in Qatar was 142 per 1000 population, compared with 97.6 in Italy, 117.6 in the United States, and 102.8 in the United Kingdom [13]. Higher COVID-19-associated mortality rates are often reported from settings where COVID-19 testing is not readily available for those who do not have severe symptoms,, thus skewing outcome assessments towards the severe end of the COVID-19’s clinical spectrum [2-4,7]. On the other hand, nearly one third of patients reported in our study were identified through screening efforts. Our lower mortality rates could therefore be in part due to higher detection of milder COVID-19 cases.

Secondly, our cohort’s demographic profile is consistent with the country’s population being largely constituted of male migrants working in the country’s numerous infrastructure projects [14]. Older age and the presence of multiple co-morbidities have consistently been associated with increased risk of severe COVID-19, need for critical care support, and mortality [7,9,15]. With a median patient age of 35 years (IQR 28-43), our patients were considerably younger than those reported in large cohorts from Lombardy Region in Italy (median 63 year, IQR 56–70), the United Kingdom (median 73 years, IQR 58–82years) and New York City (median 54 years, IQR 38–66).[5-7] Furthermore, the majority (83%) of patients in our study did not have any pre-existing chronic medical conditions.

A third factor in explaining our low COVID-19-associated mortality is the rapid escalation of the healthcare system’s capacity to accommodate the expected hike in demand for hospital beds in general, and for ICU support in particular. It has been suggested that some of the worst COVID-19-associated mortality rates have in part been the result of overwhelmed critical care resources that could not support a large influx of severely ill COVID-19 patients [3,16]. This has stimulated discussions around rationing of critical care support for COVID-19 patients, including potentially difficult decisions to withdraw resources from one patient to provide them to another [17]. On the other hand, critical care support is rarely withheld in our setting, even in cases where the prognosis appears to be unfavorable.

Deaths observed in our study have largely occurred in older patients with multiple co-morbidities. Though 85.7% of deaths occurred in those aged 55 years or above, this group constituted only 7.4% of our entire cohort. Our age group-specific mortality was 2.5% in those aged 55–64 years, and 5.4% in those aged 65 years or more. These figures are comparable with mortality rates in similar age groups in China, Italy, and the United States, but are considerably lower than those reported from the United Kingdom [6,8,9,18].

One patient in our cohort died while in a community isolation facility with asymptomatic SARS-CoV-2 infection. His rapid demise after complaining of chest pain suggests that his death was caused by myocardial infarction or pulmonary embolism. Both complications are increasingly recognized associations with COVID-19 [19,20]. An increase in out-of-hospital cardiac arrests has been observed in association with SARS-CoV-2 pandemic, including in patients with symptoms compatible with COVID-19 [21]. Moreover, 14.8% of our ICU patients and 0.3% of our non-ICU patients had evidence of myocardial injury during their hospitalization. The diagnosis of COVID-19 in patients with known or increased risk of coronary artery disease should be an opportunity to review and optimize medical therapy to reduce the risk of acute coronary events.

The majority of hospitalized patients in our study received investigational antiviral therapies. However, recent reports from large cohort and randomized clinical trials do not support the use of hydroxychloroquine, alone or in combination with azithromycin, or lopinavir-ritonavir for patients with COVID-19.[22,23] It is likely that COVID-19 management will continue to evolve as more results from ongoing clinical trials become available [24].

Our analysis showed that increasing age, male sex, higher BMI, and the presence of diabetes or chronic kidney disease are risk factors for admission to ICU. Remarkably, hypertension, chronic lung disease, and coronary artery disease, all of which are frequently reported as important predictors for severe COVID-19 in previous studies, were not independently associated with ICU admission in our setting [25]. Furthermore, our univariable analysis showed that presenting with dyspnea and cough as well as baseline blood abnormalities such as lower lymphocyte count, higher CRP and serum creatinine are associated with increased risk of admission to ICU [25,26].

Higher BMI as a risk factor for severe COVID-19 is particularly noteworthy [27]. Median BMI in our hospitalized patients was 26.8 kg/m^2^ (IQR 23.9–29.8), a reflection of the growing concern over the increasing prevalence of overweight and obesity in developing countries, along with its consequent health problems such as diabetes and cardiovascular disease [28]. In the context of COVID-19, it is important to recognize the role of overweight and obesity as a driver of severe COVID-19. Our findings help guide deployment of medical resources to better select patients for hospitalization, closer clinical monitoring, and early clinical support.

Healthcare workers represented 2.7% of cases in our report. Three (2.2%) of our healthcare workers required admission to ICU. Unlike some unfortunate reports from elsewhere, all healthcare workers in our study fully recovered within the study follow up period [29]. Risk to healthcare personnel is highest in those with prolonged direct contact with symptomatic patients, especially where personal protective equipment are either in short supply or not used appropriately [30]. Also noteworthy is that 28.2% of healthcare workers in this study were asymptomatic. Single center healthcare worker screening studies reported asymptomatic rates ranging from 12.2% to 34% [31,32]. The most efficient healthcare worker screening strategy that combines practicality with patient protection is still unclear.

Like previous reports, children in our study had a largely uneventful SARS-CoV-2 infection [33]. Likewise, only one (3.8%) out of 26 pregnant women in our report required admission to ICU. Overall outcomes were very good, conforming with existing understanding that pregnancy does not seem to be associated with increased risk of severe COVID-19 [33].

The limitations of this study include its observational nature and missing data for some variables. To address those limitations, we used multivariate analyses with multiple imputations to assess independent associations with the outcome. Despite this, our study benefits from being, to the best of our knowledge, the first to report 60-day outcomes of SARS-CoV-2, and to do so at a nationwide level.

## 5. Conclusions

In a setting of proactive SARS-CoV-2 case finding, a younger population, and low co-morbidity burden, SARS-CoV-2 was associated with low all-cause mortality. Independent risk factors for ICU admission included older age, male sex, higher BMI, and co-existing diabetes or chronic kidney disease.

## Data Availability

The datasets used and analyzed during the current study are available from the corresponding author on reasonable request.

## Acknowledgments

We would like to thank all colleagues in Hamad Medical Corporation and the Ministry of Public Health for their outstanding service and dedication.

## References

1. World Health Organization. World Health Organization. Coronavirus disease 2019 (COVID-19) Situation report – 174. Availabe online: https://www.who.int/docs/default-source/coronaviruse/situation-reports/20200712-covid-19-sitrep-174.pdf?sfvrsn=5d1c1b2c_2 (accessed on July 13, 2020)

2. Baud D.; Qi X.; Nielsen-Saines, K.; Musso, D.; Pomar, L.; Favre, G. Real estimates of mortality following COVID-19 infection. Lancet Infect Dis 2020, doi:10.1016/S1473-3099(20)30195-X.

3. Boccia, S.; Ricciardi, W.; Ioannidis, J.P.A. What Other Countries Can Learn From Italy During the COVID-19 Pandemic. JAMA Intern Med 2020, doi:10.1001/jamainternmed.2020.1447.

4. Public Health England. COVID-19: investigation and initial clinical management of possible cases. Availabe online: https://www.gov.uk/government/publications/wuhan-novel-coronavirus-initial-investigation-of-possible-cases/investigation-and-initial-clinical-management-of-possible-cases-of-wuhan-novel-coronavirus-wn-cov-infection#interim-definition-possible-cases (accessed on July 13, 2020)

5. Grasselli, G.; Zangrillo, A.; Zanella, A.; Antonelli, M.; Cabrini, L.; Castelli, A.; Cereda, D.; Coluccello, A.; Foti, G.; Fumagalli, R., et al. Baseline Characteristics and Outcomes of 1591 Patients Infected With SARS-CoV-2 Admitted to ICUs of the Lombardy Region, Italy. JAMA 2020, doi:10.1001/jama.2020.5394.

6. Docherty, A.B.; Harrison, E.M.; Green, C.A.; Hardwick, H.E.; Pius, R.; Norman, L.; Holden, K.A.; Read, J.M.; Dondelinger, F.; Carson, G., et al. Features of 20 133 UK patients in hospital with covid-19 using the ISARIC WHO Clinical Characterisation Protocol: prospective observational cohort study. BMJ 2020, 369, m1985, doi:10.1136/bmj.m1985.

7. Petrilli, C.M.; Jones, S.A.; Yang, J.; Rajagopalan, H.; O’Donnell, L.; Chernyak, Y.; Tobin, K.A.; Cerfolio, R.J.; Francois, F.; Horwitz, L.I. Factors associated with hospital admission and critical illness among 5279 people with coronavirus disease 2019 in New York City: prospective cohort study. BMJ 2020, 369, m1966, doi:10.1136/bmj.m1966.

8. Novel Coronavirus Pneumonia Emergency Response Epidemiology Team. The Epidemiological Characteristics of an Outbreak of 2019 Novel Coronavirus Diseases (COVID-19) — China, 2020. China CDC Weekly 2020, 2, 111–122, doi:10.46234/ccdcw2020.032.

9. CDC COVID Response Team. Severe Outcomes Among Patients with Coronavirus Disease 2019 (COVID-19) - United States, February 12-March 16, 2020. MMWR Morb Mortal Wkly Rep 2020, 69, 343–346, doi:10.15585/mmwr.mm6912e2.

10. Marshall, J.C.; Murthy, S.; Diaz, J.; Adhikari, N.; Angus, D.C.; Arabi, Y.M.; Baillie, K.; Bauer, M.; Berry, S.; Blackwood, B., et al. A minimal common outcome measure set for COVID-19 clinical research. Lancet Infect Dis 2020, doi:10.1016/s1473-3099(20)30483-7.

11. World Health Organization. Clinical management of COVID-19 - interim guidance 27 May 2020. Availabe online: https://apps.who.int/iris/rest/bitstreams/1278777/retrieve (accessed on July 13, 2020)

12. von Elm, E.; Altman, D.G.; Egger, M.; Pocock, S.J.; Gotzsche, P.C.; Vandenbroucke, J.P.; Initiative, S. The Strengthening the Reporting of Observational Studies in Epidemiology (STROBE) statement: guidelines for reporting observational studies. Ann Intern Med 2007, 147, 573–577, doi:10.7326/0003-4819-147-8-200710160-00010.

13. Our World in Data. Coronavirus (COVID-19) Testing. Availabe online: https://ourworldindata.org/coronavirus-testing (accessed on July 13, 2020)

14. Qatar Ministry of Development Planning Statistics. Qatar in Figures. Availabe online: https://www.mdps.gov.qa/en/statistics/Statistical%20Releases/General/QIF/Qatar_in_Figures_32_2017_EN.pdf (accessed on July 13, 2020)

15. Wu, J.T.; Leung, K.; Bushman, M.; Kishore, N.; Niehus, R.; de Salazar, P.M.; Cowling, B.J.; Lipsitch, M.; Leung, G.M. Estimating clinical severity of COVID-19 from the transmission dynamics in Wuhan, China. Nat Med 2020, 26, 506–510, doi:10.1038/s41591-020-0822-7.

16. Ji, Y.; Ma, Z.; Peppelenbosch, M.P.; Pan, Q. Potential association between COVID-19 mortality and health-care resource availability. Lancet Glob Health 2020, 8, e480, doi:10.1016/S2214-109X(20)30068-1.

17. White, D.B.; Lo, B. A Framework for Rationing Ventilators and Critical Care Beds During the COVID-19 Pandemic. JAMA 2020, doi:10.1001/jama.2020.5046.

18. Onder, G.; Rezza, G.; Brusaferro, S. Case-Fatality Rate and Characteristics of Patients Dying in Relation to COVID-19 in Italy. JAMA 2020, doi:10.1001/jama.2020.4683.

19. Shi, S.; Qin, M.; Shen, B.; Cai, Y.; Liu, T.; Yang, F.; Gong, W.; Liu, X.; Liang, J.; Zhao, Q.; et al. Association of Cardiac Injury With Mortality in Hospitalized Patients With COVID-19 in Wuhan, China. JAMA Cardiol 2020, doi:10.1001/jamacardio.2020.0950.

20. Bompard, F.; Monnier, H.; Saab, I.; Tordjman, M.; Abdoul, H.; Fournier, L.; Sanchez, O.; Lorut, C.; Chassagnon, G.; Revel, M.P. Pulmonary embolism in patients with Covid-19 pneumonia. Eur Respir J 2020, doi:10.1183/13993003.01365-2020.

21. Baldi, E.; Sechi, G.M.; Mare, C.; Canevari, F.; Brancaglione, A.; Primi, R.; Klersy, C.; Palo, A.; Contri, E.; Ronchi, V.; et al. Out-of-Hospital Cardiac Arrest during the Covid-19 Outbreak in Italy. N Engl J Med 2020, doi:10.1056/NEJMc2010418.

22. Rosenberg, E.S.; Dufort, E.M.; Udo, T.; Wilberschied, L.A.; Kumar, J.; Tesoriero, J.; Weinberg, P.; Kirkwood, J.; Muse, A.; DeHovitz, J.; et al. Association of Treatment With Hydroxychloroquine or Azithromycin With In-Hospital Mortality in Patients With COVID-19 in New York State. JAMA 2020, doi:10.1001/jama.2020.8630.

23. Cao, B.; Wang, Y.; Wen, D.; Liu, W.; Wang, J.; Fan, G.; Ruan, L.; Song, B.; Cai, Y.; Wei, M.; et al. A Trial of Lopinavir-Ritonavir in Adults Hospitalized with Severe Covid-19. N Engl J Med 2020, 382, 1787–1799, doi:10.1056/NEJMoa2001282.

24. Lythgoe, M.P.; Middleton, P. Ongoing Clinical Trials for the Management of the COVID-19 Pandemic. Trends Pharmacol Sci 2020, 41, 363–382, doi:10.1016/j.tips.2020.03.006.

25. Zheng, Z.; Peng, F.; Xu, B.; Zhao, J.; Liu, H.; Peng, J.; Li, Q.; Jiang, C.; Zhou, Y.; Liu, S.; et al. Risk factors of critical & mortal COVID-19 cases: A systematic literature review and meta-analysis. J Infect 2020, 10.1016/j.jinf.2020.04.021, doi:10.1016/j.jinf.2020.04.021.

26. Jain, V.; Yuan, J.M. Predictive symptoms and comorbidities for severe COVID-19 and intensive care unit admission: a systematic review and meta-analysis. Int J Public Health 2020, doi:10.1007/s00038-020-01390-7.

27. Simonnet, A.; Chetboun, M.; Poissy, J.; Raverdy, V.; Noulette, J.; Duhamel, A.; Labreuche, J.; Mathieu, D.; Pattou, F.; Jourdain, M.; et al. High Prevalence of Obesity in Severe Acute Respiratory Syndrome Coronavirus-2 (SARS-CoV-2) Requiring Invasive Mechanical Ventilation. Obesity (Silver Spring) 2020, doi:10.1002/oby.22831.

28. Yoon, K.H.; Lee, J.H.; Kim, J.W.; Cho, J.H.; Choi, Y.H.; Ko, S.H.; Zimmet, P.; Son, H.Y. Epidemic obesity and type 2 diabetes in Asia. Lancet 2006, 368, 1681–1688, doi:10.1016/S0140-6736(06)69703-1.

29. Lapolla, P.; Mingoli, A.; Lee, R. Deaths from COVID-19 in healthcare workers in Italy-What can we learn? Infect Control Hosp Epidemiol 2020, doi:10.1017/ice.2020.241.

30. Wang, J.; Zhou, M.; Liu, F. Reasons for healthcare workers becoming infected with novel coronavirus disease 2019 (COVID-19) in China. J Hosp Infect 2020, 105, 100–101, doi:10.1016/j.jhin.2020.03.002.

31. Khalil, A.; Hill, R.; Ladhani, S.; Pattisson, K.; O’Brien, P. COVID-19 screening of health-care workers in a London maternity hospital. Lancet Infect Dis 2020, doi:10.1016/S1473-3099(20)30403-5.

32. Lombardi, A.; Consonni, D.; Carugno, M.; Bozzi, G.; Mangioni, D.; Muscatello, A.; Castelli, V.; Palomba, E.; Cantù, A.P.; Ceriotti, F.; et al. Characteristics of 1,573 healthcare workers who underwent nasopharyngeal swab for SARS-CoV-2 in Milano, Lombardy, Italy. Clin Microbiol Infect 2020, doi:10.1016/j.cmi.2020.06.013.

33. Zimmermann, P.; Curtis, N. COVID-19 in Children, Pregnancy and Neonates: A Review of Epidemiologic and Clinical Features. Pediatr Infect Dis J 2020, 39, 469–477, doi:10.1097/INF.0000000000002700.

